# Assessment of level of depression and associated factors among covid-19 recovered patients: a cross sectional study

**DOI:** 10.1101/2022.11.14.22282128

**Authors:** Mst.Khadeza Khatun, Nasreen Farhana

## Abstract

**Objectives:** The Corona Virus Disease-2019 (COVID-19) pandemic has psychological consequences such as increased risk of depression, anxiety, and stress problems, exacerbating human health disparities. This study aimed to analyze depression and its causes in COVID-19-recovered patients in Bangladesh.

**Method:** A cross-sectional study was conducted on COVID-19 recovered patients, who attended for follow-up after 14 days to 3 months at Dhaka Medical College Hospital (DMCH) and Dhaka North City Corporation Hospital (DNCCH), Dhaka, Bangladesh from 1st January to 31st December, 2021. Respondents were face-to-face interviewed with a semi-structured questionnaire after written agreement. The Patient Health Questionnaire (PHQ-9) was used to assess respondents’ depression, and data were analyzed using SPSS version-23, with p < 0.05 indicating statistical significance.

**Results:** A total of 325 COVID-19 recovered patients aged from 15 to 65 years (mean 44.34 ±13.87 years) of age were included in this study, highest 23.1% of them belonged to 46-55 years, and majority (61.5%) of them were male. There were 69.5% of respondents had no signs of depression while 31% of them had with 26.7 % being mildly depressed, 2.5 % being extremely depressed, and 1.2 % being severely depressed. Diabetes mellitus, hospitalization duration, social distancing, the social media post on COVID-19, loss of employment, family damage, and fear of re-infection were significantly associated with depression level of respondents.

**Conclusion:** This study gives us a glimpse into the psychological health of COVID-19 recovered patients, and its findings highlight the imperative of alleviating their psychological anguish in Bangladesh.

## Introduction

Since the dawn of 2020, the world has faced a pandemic caused by SARS-CoV-2, the highly contagious COVID-19 (1). Bangladesh has been suffering from this highly transmissible disease since March 2020 (2). The rapid spread of the disease in a wide variety of people makes them subject to varied degrees of panic, which makes it difficult to treat and rehabilitate them (3). According to earlier studies, viral respiratory infections are linked to chronic and acute psychological effects among the survivors including posttraumatic stress, insomnia, depression, anxiety and even suicidality. After discharge, many of these patients’ mental disturbances persisted and continued for a long time (4, 5).

The COVID-19 pandemic has a significant psychological impact on healthy populations, with an increase in depression, perceived stress, post-traumatic stress, and insomnia being reported (6). The COVID-19 pandemic situation is fast changing with worldwide case counts of crisis, suicide, domestic violence, mental disorder, anxiety, depressive disorders (7). To combat the spread of COVID-19, the Bangladesh government has implemented a number of measures, including lockdown, social distancing, self-isolation, and quarantine (8). Besides, recent findings have demonstrated prevailing acute psychiatric symptoms among patients while being treated in isolation, lack of interaction with friends, family, or loved ones, admitted to intensive care units (ICUs) and required mechanical ventilation (9). Extreme fear, when accompanied with social and economic implications (e.g., job loss, reduced earnings, relationship problems), has the potential to encourage individuals to engage in irrational thinking, which can lead to psychological discomfort (10).

In prior lethal viral epidemics, fear of death, loneliness, boredom, anxiety, sadness, social isolation, contagion, and (in severe situations) thoughts of suicide might result in long-term psychological repercussions among the general population (11). In the present COVID-19 pandemic, depression and anxiety problems are more activated. A recent systematic review reported that the pooled prevalence of depression and anxiety among COVID-19 patients was 45% and 47%, respectively (12).

Patients with COVID-19 have different degree of psychological pain such as anxiety and depression, which may related to their prognosis worse by negatively affect the patients’ immunity. As a result, clinically significant depression could have serious consequences for quality of life (13). Despite its increasing significance, present estimates on the prevalence of psychological distress, such as anxiety and depressive symptoms, among COVID-19 patients are uncertain (12).

In the aftermath of a natural disaster, a high rate of mental health suffering (i.e, a prevalence rate of 65% depression, much higher than rates reported elsewhere) was previously reported, and appears to reflect the vulnerability of the Bangladeshi population to mental health suffering in a pandemic situation such as COVID-19 (14). During pandemics, public health officials and the media are more concerned with the biological and physical consequences of the outbreak than with mental health difficulties. With an increasing number of reports indicating that the COVID-19 outbreak is causing an increase in mental health burden, there have been greater calls for steps to improve public mental health support for the public (15).

To date, the scant reported findings on COVID-19 have been conducted regarding the prevalence and associated factors of depression among COVID-19 recovered patients in Bangladesh. Therefore, this study aimed to comprehensively assess the level of depression and explore the factors associated with depressive symptoms 3 months after recovery from COVID-19 infection.

## Methods Study

### design

This cross-sectional study was conducted among COVID-19 recovered patients, who came for follow up after 14 days up to 3 month at post COVID unit of Dhaka Medical College Hospital (DMCH) and Dhaka North City Corporation Hospital (DNCCH), Dhaka, Bangladesh from 1st January to 31st December, 2021. Patients with known case of depressive illness and other psychiatric disorder were excluded from this study.

### Sampling and sample size

A non-probability convenience sampling method was used and to estimate the required sample size, the prevalence for depression was set as 47.2% (16). So the calculated sample size was 382, but because of the unfavorable COVID-19 scenario, it was possible to conduct interviews with 325 respondents during the designated data collection period.

### Ethical considerations

Ethical approval for the study was granted by the Institutional Review Board (IRB) of National Institute of Preventive and Social Medicine (NIPSOM) with memo no. NIPSOM/IRB/2021/18 dated 13.12.2021.

### Data collection

According to the specific objectives and after the pretest observation, a semi structured questionnaire was developed in English and then translated to Bengali using the selected variables. The respondents were explained the title and purpose of the study, their anonymity, and voluntary participation of the study by ensuring the privacy and confidentiality.

### Data collection instrument

After informed written consent, the respondents were interviewed by face to face with the questionnaire examining socio-demographic variables and the factors that causing depression among them. Questionnaire was developed by using Patient Health Questionnaire (PHQ-9).

### Level of depression

PHQ-9 has nine different questions that assess the depressive symptoms of the respondents. The total score ranges from 0 to 27 points, where each question is scored from 0 to 3 depending on the answer: 0 (not at all), 1 (several days), 2 (half of the days), and 3 (nearly every day). To determine the state of depression, the total score was divided into four score: cumulative scores <10 indicate no depression, 10– 15 indicate mild depression, 16–21 indicate moderate depression and 22–27 indicate severe depression (17).

### Statistical analysis

Data processing and analysis was done using Statistical Package for Social Sciences (SPSS) version23 according to objectives and variables. Frequency, percentage, mean and standard deviation (SD) were used for descriptive statistics and Chi square (X^2^) test were carried out to assess the association of qualitative data with 95% confidence interval (CI), and P<0.05 was considered statistically significant. Data were presented through tables and figures.

## Results

### The socio demographic characteristics of the respondents

This study comprised a total of 325 respondents, whose ages ranged from 15 to 65 years. The respondents’ mean age was 44.34 years, with SD of ±13.87 years, and the highest 23.1% were between 46 and 55 years old. The majority of respondents (61.5%) were men; 75.7% were married; 92% were Muslims; 21.2% were graduates; 31.4% were business owners; 81.5% belonged to nuclear families; 61.5% resided in urban areas; 89.2% of them had only one family member who was financially contributing; 37% of respondents did not complete vaccination against COVID-19, and hypertension was found to be the most prevalent chronic disease (Table 1).

**Table-1:**
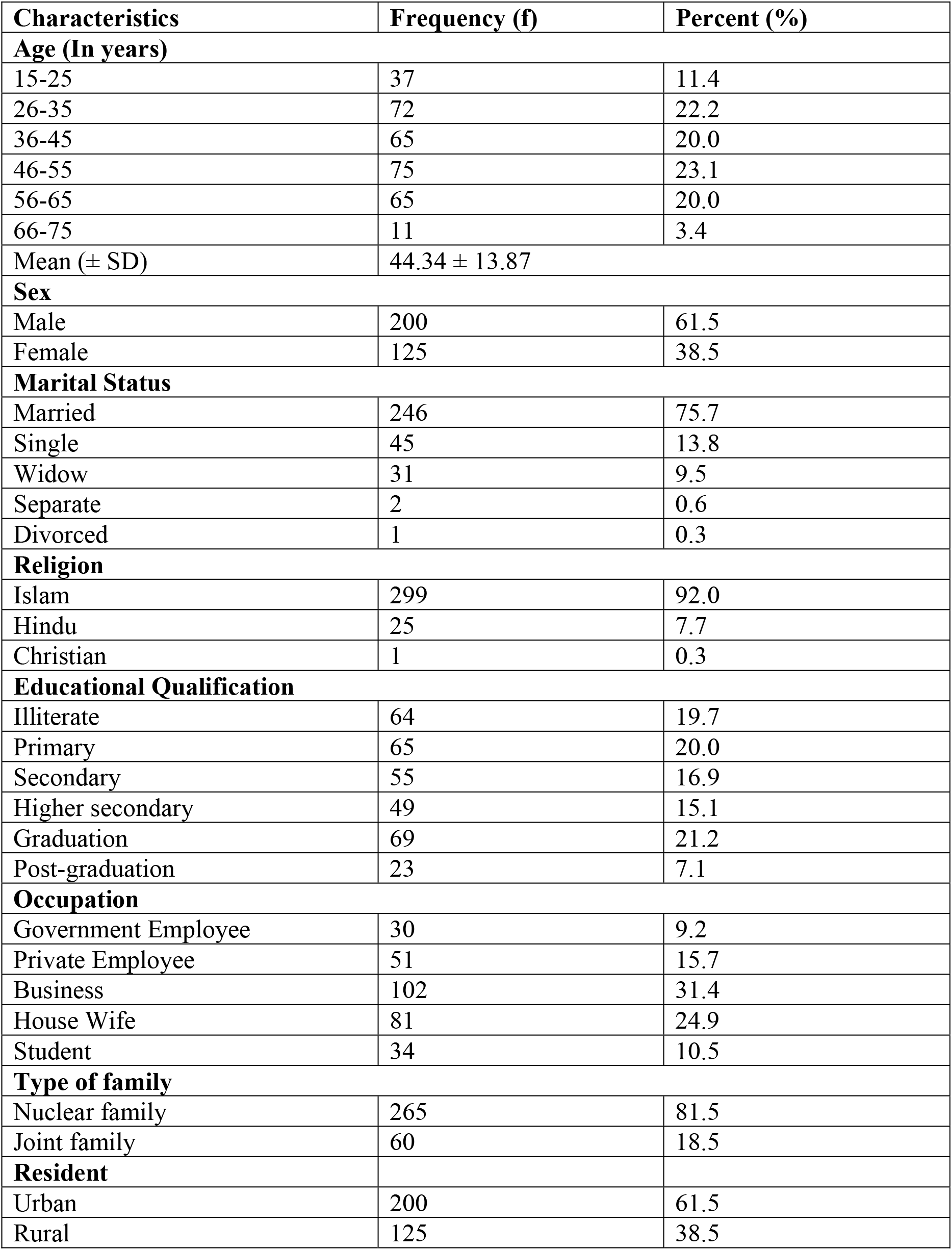
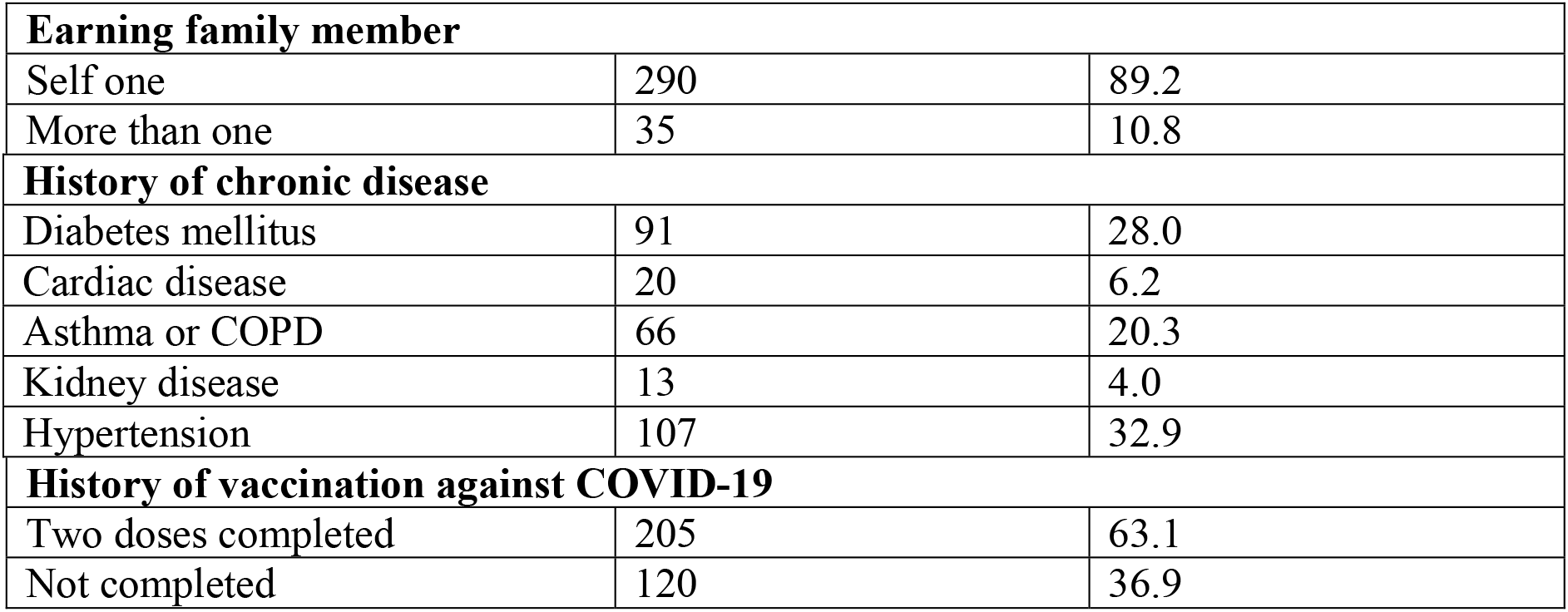
Socio-demographic characteristics of the respondents.

### Distribution of the conditions of the COVID-19 recovered patients

One-fourth of 325 respondents (25.2%) had personal life interruptions and 78.2% had physical symptoms after COVID-19, primarily weakness (57%) and cough (31%). Of them, 4.3% were taking anti-depressants, 1.2% had a family history of depression, 12% were hospitalized for more than 21 days, 10.8% needed ICU, and 12.3% were distressed due to social distancing. 10.2% were affected by social media posts about COVID-19, 13% felt harmful to the family, 2.8% feared losing their jobs, 6.2% feared re-infection with COVID-19, and 29% had relatives or acquaintances who died from COVID-19 (Table 2).

**Table 2:** Distribution of the conditions of the COVID-19 recovered patients (n=325)

| **Variable** | **Frequency** | **Percent** |
| --- | --- | --- |
| **Interrupting personal life** | 82 | 25.2 |
| **Physical symptoms after recovery** | 254 | 78.2 |
| **Physical symptoms** |  |  |
| Dyspnea | 63 | 19.4 |
| Weakness | 184 | 56.6 |
| Cough | 101 | 31.1 |
| Fatigue | 85 | 26.2 |
| Muscle pain | 30 | 9.2 |
| Palpitation | 45 | 13.8 |
| **Taking anti- depressant drug** | 14 | 4.3 |
| **Family history of depression** | 4 | 1.2 |
| **Duration of hospitalization** |  |  |
| 1-7 days | 183 | 56.3 |
| 7-14 days | 61 | 18.8 |
| 14-21 days | 33 | 10.2 |
| More than 21 days | 39 | 12.0 |
| **Need ICU** | 35 | 10.8 |
| **Mentally distress due to social distancing** | 40 | 12.3 |
| **Affected by the post on social media about COVID 19** | 33 | 10.2 |
| **Do you feel that you can be harmful to your family** | 42 | 12.9 |
| **Fear of losing job** | 9 | 2.8 |
| **Fear of re-infection with COVID-19** | 20 | 6.2 |
| **Relative or acquaintances died from COVID 19** | 93 | 28.6 |

### Prevalence of depression

The prevalence of depression was assessed by PHQ-9 scale comprising 9 items (Table 3) showed that about one third of them (31%) had symptoms of depression (Figure 1), whereas 87.8% of them were mildly depressed, 8.1% were moderately depressed, and 4.0% were severely depressed (Table 4).

**Table 3:** Depression related questions according to patient health questionnaire PHQ-9scale (9 items) (n=325)

| **Attribute** | **Not at all** | **Several day** | **More than half day** | **Nearly everyday** |
| --- | --- | --- | --- | --- |
| Little interest or pleasure in doing things | 244 (75.1%) | 19 (5.8%) | 27 (8.3%) | 35 (10.8%) |
| Feeling down or hopeless | 226 (69.5%) | 51 (15.7%) | 19 (5.8%) | 29 (8.9%) |
| Trouble falling or staying sleeping too much | 139 (42.8%) | 74 (22.8%) | 8 (2.5%) | 104 (32%) |
| Feeling tired or having little energy | 103 (31.7%) | 81 (24.9%) | 6 (1.8%) | 135 (41.5%) |
| Poor appetite or over eating | 149 (45.8%) | 48 (14.8%) | 6 (1.8%) | 122 (37.5%) |
| Feeling bad about yourself | 292 (89.8%) | 11 (3.4%) | 18 (5.5%) | 4 (1.2%) |
| Trouble concentrating | 283 (87.1%) | 25 (7.7%) | 3 (0.9%) | 14 (4.3%) |
| Moving or speaking slowly | 271 (83.4%) | 44 (13.5%) | 6 (1.8%) | 4 (1.2%) |
| Thoughts would be better dead | 307 (94.5%) | 6 (1.8%) | 3 (0.9%) | 9 (2.8%) |

**Table 4:** Distribution of respondents according to level of depression (n=99)

| **Level of depression** | **Frequency (f)** | **Percent (%)** |
| --- | --- | --- |
| Mild depression | 87 | 87.9 |
| Moderate depression | 8 | 8.1 |
| Severe depression | 4 | 4.0 |

**Figure 1:**
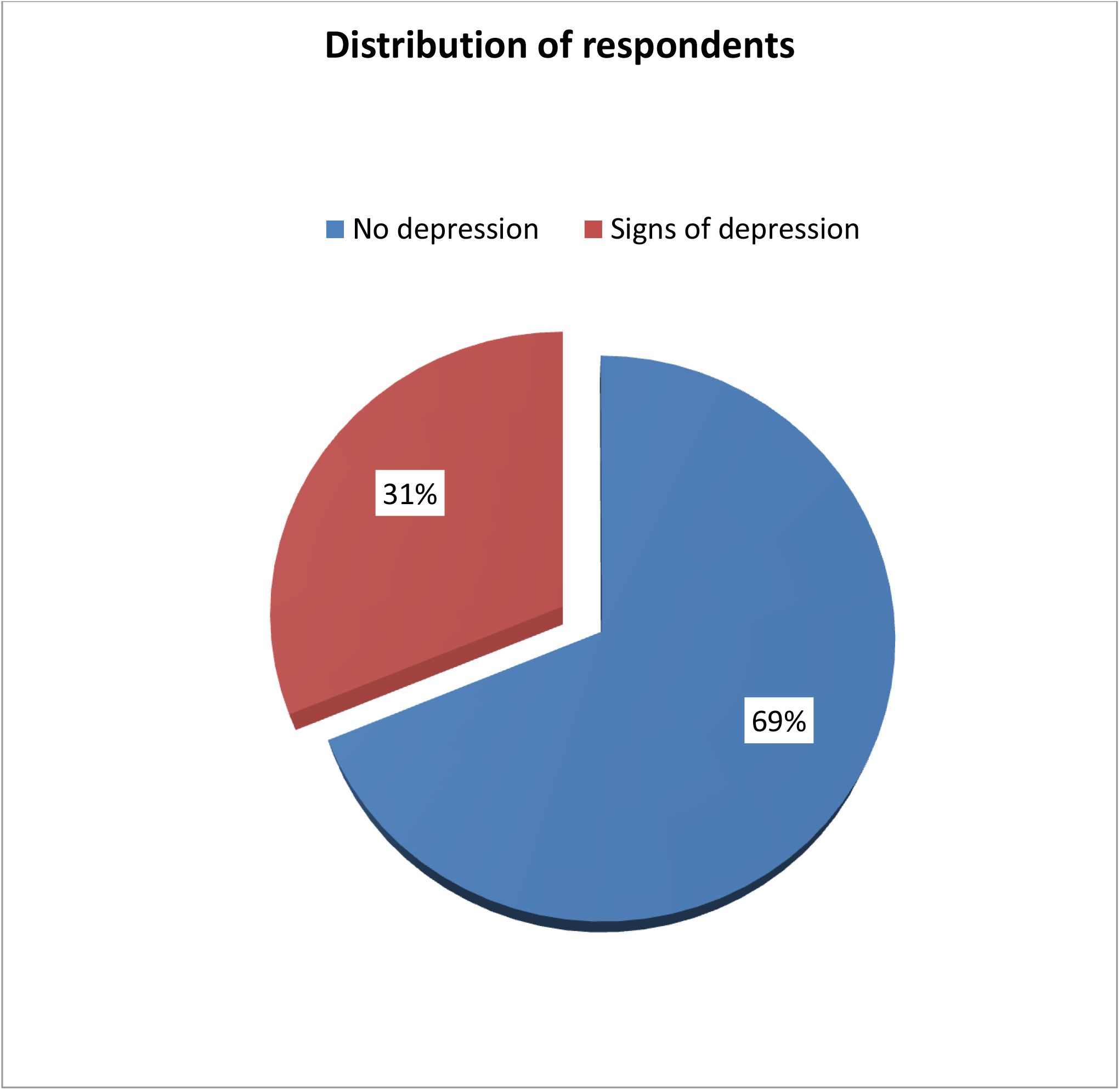
Distribution of respondents according to signs of depression (n=325)

### Association of prevalence of depression with the variables of the respondent

The variables of educational qualification, diabetes mellitus history, duration of hospitalization, distress due to social distancing, affected by the post on social media about COVID 19, fear of losing job, feeling harmful to family members, fear of re-infection, taking anti-depressant drug, interrupting personal life were significantly associated with the prevalence of depression (P < 0.05) (Table 5).

**Table 5:**
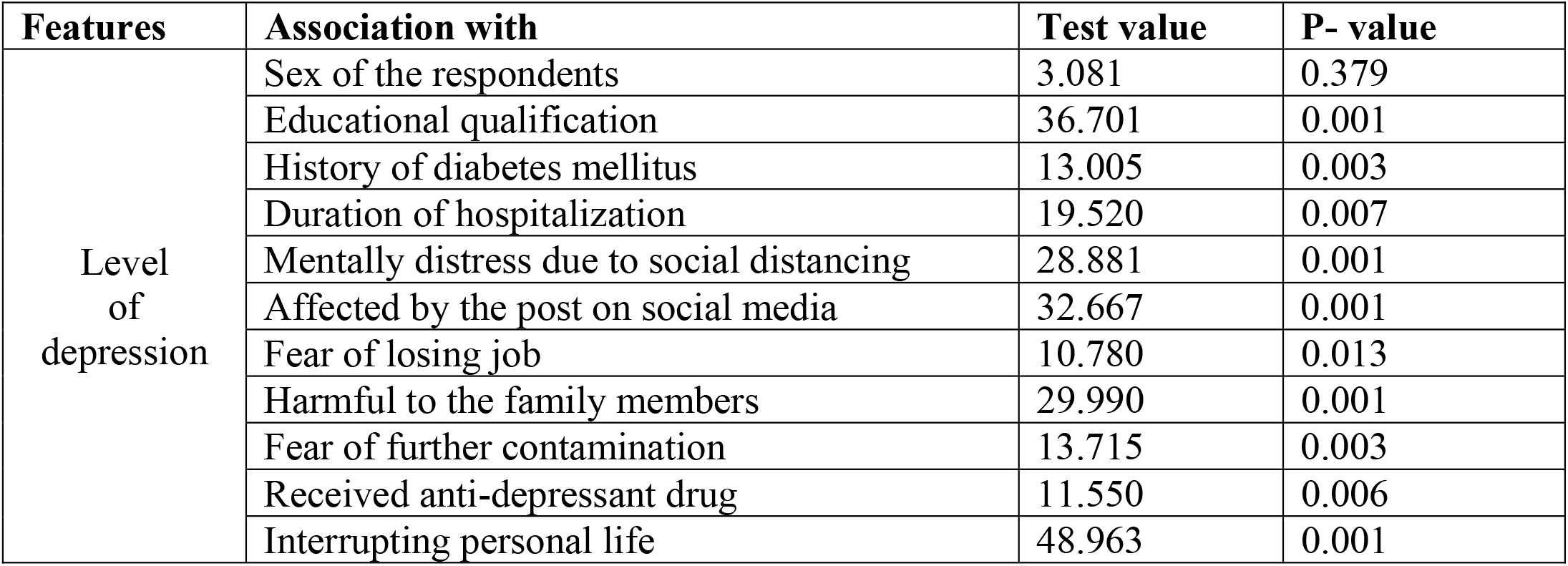
Association of level of depression with the variables of the respondent (n=325)

| **Features** | **Association with** | **Test value** | **P- value** |
| --- | --- | --- | --- |
| Level<br>of<br>depression | Sex of the respondents | 3.081 | 0.379 |
|  | Educational qualification | 36.701 | 0.001 |
|  | History of diabetes mellitus | 13.005 | 0.003 |
|  | Duration of hospitalization | 19.520 | 0.007 |
|  | Mentally distress due to social distancing | 28.881 | 0.001 |
|  | Affected by the post on social media | 32.667 | 0.001 |
|  | Fear of losing job | 10.780 | 0.013 |
|  | Harmful to the family members | 29.990 | 0.001 |
|  | Fear of further contamination | 13.715 | 0.003 |
|  | Received anti-depressant drug | 11.550 | 0.006 |
|  | Interrupting personal life | 48.963 | 0.001 |

## Discussion

This study found that among 325 respondents, 31% had depressed symptoms, with 26.8% exhibiting mild symptoms, 2.5% exhibiting moderate symptoms, and 1.2% exhibiting severe symptoms. Several studies were undertaken in Bangladesh following the commencement of COVID-19 to assess mental health, including depression. A cross sectional study was conducted at the initial stage after the COVID-19 outbreak (from June 1 to June 10, 2020) among 1146 Bangladeshi participants by Zubayer, et al., (16) in which 47.2% of participants had depression, with 16.2% were mildly depressed, 20.4% moderately depressed, 6.5% were severely depressed which was higher than this study. Another study conducted by Das, et al., (17) among 672 Bangladeshi people aged between 15 and 65 years from 15 April to 10 may 2020 where the prevalence of depression are 38% with 24% were mildly depressed, 11% moderately depressed and 3% were severely depressed. Another study conducted among Bangladeshi students during COVID-19 pandemic reported the prevalence rates of depression to be 46.9% (18). The increased prevalence of depression could be attributed to the virus’s ongoing transmission, the increasing number of new cases, the death of a loved one, and the fear of infection during the early stages of the COVID-19 outbreak, when individuals were challenged by mandatory quarantine, unexpected unemployment, and uncertainty associated with the outbreak (16,18).

According to certain research, depressive symptoms were recorded in lower prevalence rates than the present study like 16.5% of the Chinese population (19), 11.4% of Japanese people (20), 17.3 % of Italians (3), 3.7% of Portuguese people (21), and 26% of Iranian people (22). Prevalence rates varied between studies, which could be explained by differences in government preparedness, the availability of medical supplies and facilities, the effective communication of information about COVID-19, or international/cultural variances affecting the psychological health of the general public (18).

According to the results of this study, the highest 23.1% of the respondents were between the ages of 46 to 55 years. It is because the risk of COVID-19-related illness and mortality increases with age (8). In some studies, individuals under the age of 45 had more adverse psychological symptoms during the pandemic (23, 24). This is consistent with our findings that about 54% of the participants were below 45 years of age. This finding could be explained in part by their role as family caregivers, who provide financial and emotional support to children and the elderly. Among the respondents, 61.5% were male, although females have been linked to mental health issues, but there were no significant sex differences (p=0.379) in regard to depression in the current investigation (18, 25). Because of the COVID-19 condition, sex disparities in this study may be negated.

COVID-19 is highly contagious and can be passed from person to person (26). Therefore, Individuals fear either of having COVID-19 themselves or of becoming asymptomatic carriers who spread the disease unknowingly to family and friends, contributing to psychiatric symptoms (21, 26). In line with previous studies, the present study also showed that fear of infection is significantly associated with depression (P=0.001) (23, 27). The death of a beloved leads to psychological problems, such as depression, is supported by the findings of the present study (26).

The present study showed that daily exposure to COVID-19 related news was significantly associated with overall mental health problems. Previous studies showed that people who exposed COVID-19 related news were more likely to develop psychiatric symptoms (23, 28). Moreover, symptoms of depression and anxiety among COVID-19 inpatients could increase due to uncertainty about the prognosis of the disease and the experiencing the adverse outcomes (16). Furthermore, side effects of COVID-19 medication and physical discomfort may also promote psychiatric problems among COVID-19 inpatients (28). This high prevalence of anxiety and depressive symptoms among Bangladeshi COVID-19 inpatients could be because of the incompetent healthcare system (8), shortage of beds, ICU and ventilator (16), treatment-related negligence in the healthcare facilities (29), less social interaction and rampant circulation of misinformation on social and conventional media (30).

In this study the result showed that 78.2% having physical symptoms after recovery from COVID-19 and weakness (56.6%) was the most prevalent feature. However, a prospective cohort study among the Bangladeshi population reported that the incidence of the post-COVID-19 syndrome was 46% and other features included persistent cough (8.5%), post-exertional dyspnea (7%), headache (3.4%), vertigo (2.3%), and sleep-related disorders (5.9%), which were lower than in the present study (31). COVID-19 can cause acute respiratory syndrome with consequent release of pro-inflammatory cytokines, including interleukin (IL)-1β and IL-6 from the respiratory tract. These cytokines were commonly found to be increased in major depressive disorder (32)

Due to the COVID-19 epidemic, we have had to make some adjustments to our regular schedule. Because of the substantial changes in everyday lives, in this study, 25.2% had suffered from interruption of regular personal life, which has led to concerns with other studies on mental health (3, 16, 23).

The Pearson Chi-Square test was used in this study to evaluate the association between depression and sociodemographic characteristics with the factors related COVID-19 recovered patients. According to the results, education qualification (P=0.001), co-morbidity diabetes mellitus (p=0.001), received anti-depressant drug (p=0.006), having fear of further contamination (p=0.003), respondents who felt that they were harmful to her family member (p=0.001), having fear of losing job (p=0.013), respondents affected by the social media about COVID 19 (p=0.001), respondents are mentally distress due to social distancing (p=0.001), duration of hospitalization (p=0.007) were statistically significant with depression (P=0.001), which is consistent with other researches (12,16, 33). The current study highlights the necessity for reducing these psychological suffering in Bangladesh. Appropriate supportive programmes and interventional approaches would be addressed in Bangladesh during COVID-19 pandemic.

## Limitations

This study has a few limitations. First, data was collected conveniently from the patient during the COVID-19 recovery period which could result in recall and selection bias. Second, the risk factors of depression were not able to analyze associated with the level of depression. Third, all the respondents in this study were sampled from two selected post-COVID units in Dhaka city, so the generalization of this study was indeterminate and might not present the whole country’s scenario during the COVID-19 pandemic.

## Conclusion

The state of depression impacts on COVID-19 recovered patients is undeniable. The study found that a significant portion of respondents reported mental health problems, with different levels of severity of depression. Most of the respondents were concerned about the presence of COVID-19-related symptoms, fear of losing jobs; fear of re-infection, distress due to social distancing, etc. provoke individuals with depression. The findings of the study suggest the need for more targeted measures like health education intervention for the people infected with COVID-19 in Bangladesh to accelerate progress in reducing the incidence of depression.

## Data Availability

All data produced in the present study are available upon reasonable request to the authors

## Data availability statement

The original contributions presented in the study are included in the article/supplementary material; further inquiries can be directed to the corresponding author/s.

## Ethics statement

Ethical approval for the study was granted by the Institutional Review Board (IRB) of National Institute of Preventive and Social Medicine (NIPSOM) (No.: NIPSOM/IRB/2021/18/dated-13.12.2021). Written informed consent for participation was required for this study in accordance with the national legislation and the institutional requirements.

## Author contributions

MKK was involved in setting up the study, data collection, and data analysis. NF drafted the manuscript. All of the authors made comments on the different versions, added to the article, and approved the version that was sent in.

## Acknowledgments

The authors would like to express their gratitude to all who participated and filled out the study.

## Abbreviations

COVID-19: Corona Virus Disease-2019
PHQ-9: Patient Health Questionnaire
SD: Standard Deviation
SPSS: Statistical Package for the Social Sciences

## References

(1) World Health Organization (WHO). Coronavirus (2020c). https://www.who.int/health-topics/coronavirus#tab=tab_3 [Accessed 15 November, 2020].

(2) Director General of Health Services (DGHS). Corona Virus Info (2020). https://corona.gov.bd/ [Accessed 13 November, 2020].

(3) Liu S, Yang L, Zhang C, Xiang YT, Liu Z, Hu S, et al. Online mental health services in China during the COVID-19 outbreak. The lancet. Psychiatry (2020) 7(4), e17–e18. https://doi.org/10.1016/S2215-0366(20)30077-8

(4) Kim HC, Yoo SY, Lee BH, Lee SH, Shin HS. Psychiatric Findings in Suspected and Confirmed Middle East Respiratory Syndrome Patients Quarantined in Hospital: A Retrospective Chart Analysis. Psychiatry Investig (2018)15(4):355–60. doi: 10.30773/pi.2017.10.25.1.

(5) Lam MH, Wing YK, Yu MW, Leung CM, Ma RC, Kong AP, et al. Mental morbidities and chronic fatigue in severe acute respiratory syndrome survivors: long-term follow-up. Arch Intern Med (2009) Dec 14;169(22):2142–7. https://doi.10.1001/archinternmed.2009.384.

(6) Rajkumar RP. COVID-19 and mental health: A review of the existing literature. Asian J Psychiatr (2020) 52:102066. https://doi.10.1016/j.ajp.2020.102066.

(7) Sifat RI. Impact of the COVID-19 pandemic on domestic violence in Bangladesh. Asian J Psychiatr (2020) 53:102393. https://doi.10.1016/j.ajp.2020.102393.

(8) Anwar S, Nasrullah M, Hosen MJ. COVID-19 and Bangladesh: Challenges and How to Address Them. Front Public Health (2020) 8:154. https://doi.10.3389/fpubh.2020.00154.

(9) Li W, Yang Y, Liu ZH, Zhao YJ, Zhang Q, Zhang L, et al. Progression of Mental Health Services during the COVID-19 Outbreak in China. Int J Biol Sci (2020)15;16(10):1732–8. https://doi.10.7150/ijbs.45120.

(10) Pakpour AH, Griffiths MD. The fear of COVID-19 and its role in preventive behaviors. J Concurr Disord. (2020)2: 58–63. http://irep.ntu.ac.uk/id/eprint/39561.

(11) Mak IW, Chu CM, Pan PC, Yiu MG, Chan VL. Long-term psychiatric morbidities among SARS survivors. Gen Hosp Psychiatry. (2009) 31(4):318–26. https://doi.10.1016/j.genhosppsych.2009.03.001.

(12) Deng J, Zhou F, Hou W, Silver Z, Wong CY, Chang O, et al. The prevalence of depression, anxiety, and sleep disturbances in COVID-19 patients: a meta-analysis. Ann N Y Acad Sci. (2021) 1486(1):90–111. https://doi.10.1111/nyas.14506.

(13) Renaud-Charest O, Lui LMW, Eskander S, Ceban F, Ho R, Di Vincenzo JD, et al. Onset and frequency of depression in post-COVID-19 syndrome: A systematic review. J Psychiatr Res. (2021) 144:129–37. https://doi.10.1016/j.jpsychires.2021.09.054.

(14) Mamun MA, Hossain MS, Griffiths MD. Mental Health Problems and associated Predictors among Bangladeshi Students. Int J Ment Health Addiction (2022) 20, 657–71. https://doi.org/10.1007/s11469-019-00144-8.

(15) Ho CS, Chee CY, Ho RC. Mental Health Strategies to Combat the Psychological Impact of Coronavirus Disease 2019 (COVID-19) Beyond Paranoia and Panic. Ann Acad Med Singap.(2020) 16;49(3):155-60. PMID: 32200399.

(16) Zubayer AA, Rahman ME, Islam MB, Babu SZD, Rahman QM, Bhuiyan MRAM, et al. Psychological states of Bangladeshi people four months after the COVID-19 pandemic: An online survey. Heliyon. (2020) 6(9):e05057. https://doi.10.1016/j.heliyon.2020.e05057.

(17) Das R, Hasan MR, Daria S, Islam MR. Impact of COVID-19 pandemic on mental health among general Bangladeshi population: a cross-sectional study. BMJ Open. (2021) 9;11(4):e045727. https://doi.10.1136/bmjopen-2020-045727.

(18) Xiong J, Lipsitz O, Nasri F, Lui LMW, Gill H, Phan L, et al. Impact of COVID-19 pandemic on mental health in the general population: A systematic review. J Affect Disord. (2020) 1;277:55-64. https://doi.10.1016/j.jad.2020.08.001.

(19) Dai LL, Wang X, Jiang TC, Li PF, Wang Y, Wu SJ, et al. Anxiety and depressive symptoms among COVID-19 patients in Jianghan Fangcang Shelter Hospital in Wuhan, China. PLoS One. (2020) 28;15(8):e0238416. https://doi.10.1371/journal.pone.0238416.

(20) Ueda M, Stickley A, Sueki H, Matsubayashi T. Mental health status of the general population in Japan during the COVID-19 pandemic. Psychiatry Clin Neurosci. (2020) 74(9):505–6. https://doi.10.1111/pcn.13105.

(21) Newby JM, O’Moore K, Tang S, Christensen H, Faasse K. Acute mental health responses during the COVID-19 pandemic in Australia. PLOS ONE. (2020) 15(7): e0236562. https://doi.org/10.1371/journal.pone.0236562

(22) Vahedian-Azimi A, Moayed MS, Rahimibashar F, Shojaei S, Ashtari S, Pourhoseingholi MA. Comparison of the severity of psychological distress among four groups of an Iranian population regarding COVID-19 pandemic. BMC Psychiatry. (2020) 8;20(1):402. https://doi.10.1186/s12888-020-02804-9.

(23) Gao J, Zheng P, Jia Y, Chen H, Mao Y, Chen S, et al. Mental health problems and social media exposure during COVID-19 outbreak. PLoS ONE (2020) 15(4): e0231924. https://doi.org/10.1371/journal.pone.0231924

(24) Huang Y, Zhao N. Generalized anxiety disorder, depressive symptoms and sleep quality during COVID-19 outbreak in China: a web-based cross-sectional survey. Psychiatry Res. (2020) 288:112954. https://doi.10.1016/j.psychres.2020.112954.

(25) Moghanibashi-Mansourieh A. Assessing the anxiety level of Iranian general population during COVID-19 outbreak. Asian J Psychiatr. 2020 Jun;51:102076. https://doi.10.1016/j.ajp.2020.102076.

(26) Wang, C., Pan, R., Wan, X., Tan, Y., Xu, L., Mclntyre, R. S. et al. A longitudinal study on the mental health of general population during the COVID-19 epidemic in China. Brain Behav Immun. (2020) 87:40–8. https://doi.10.1016/j.bbi.2020.04.028.

(27) Zolotov Y, Reznik A, Bender S, Isralowitz R. COVID-19 Fear, Mental Health, and Substance Use Among Israeli University Students. Int J Ment Health Addict. (2022) 20(1):230–6. https://doi.10.1007/s11469-020-00351-8.

(28) Islam, M.S., Sujon, M. S. H., Tasnim, R., Sikder, M. T., Potenza, M. N. and Os, J. V. Psychological responses during the COVID-19 outbreak among university students in Bangladesh. PLoS ONE. (2020) 15. Pp, e0245083. https://doi.10.1371/journal.pone.0245083.

(29) Al-Zaman MS. Healthcare Crisis in Bangladesh during the COVID-19 Pandemic. Am J Trop Med Hyg. 2020 Oct;103(4):1357–1359. https://doi.10.4269/ajtmh.20-0826.

(30) Zarocostas J. How to fight an infodemic. Lancet. 2020 Feb 29;395(10225):676. https://doi.10.1016/S0140-6736(20)30461-X.

(31) Mahmud S, Mohsin M, Khan IA, Mian AU, Zaman MA. Knowledge, beliefs, attitudes and perceived risk about COVID-19 vaccine and determinants of COVID-19 vaccine acceptance in Bangladesh. PLOS ONE. (2021) 16(9): e0257096. https://doi.org/10.1371/journal.pone.0257096

(32) Conti P, Ronconi G, Caraffa A, Gallenga CE, Ross R, Frydas I, et al. Induction of pro-inflammatory cytokines (IL-1 and IL-6) and lung inflammation by Coronavirus-19 (COVID-19 or SARS-CoV-2): anti-inflammatory strategies. J Biol Regul Homeost Agents. (2020) 34(2):327–331. https://doi.10.23812/CONTI-E.

(33) Renaud-Charest O, Lui LMW, Eskander S, Ceban F, Ho R, Di Vincenzo JD, et al. Onset and frequency of depression in post-COVID-19 syndrome: A systematic review. J Psychiatr Res. 2021 Dec;144:129–137. https://doi.10.1016/j.jpsychires.2021.09.054.

